# Children with familial hypercholesterolemia display changes in LDL and HDL function: a cross-sectional study

**DOI:** 10.1101/2020.11.11.20214247

**Authors:** Jacob J. Christensen, Ingunn Narverud, Maija Ruuth, Martin Heier, Matti Jauhiainen, Stine M. Ulven, Martin P. Bogsrud, Petri T. Kovanen, Bente Halvorsen, Michael N. Oda, Cecilie Wium, Kjetil Retterstøl, Katariina Öörni, Kirsten B. Holven

## Abstract

**Background:** The functional status of lipoprotein particles contributes to atherogenesis. The tendency of plasma LDL particles to aggregate and the ability of HDL particles to induce and mediate reverse cholesterol transport associate with high and low risk for cardiovascular disease in adult patients, respectively. However, it is unknown whether children with familial hypercholesterolemia (FH) display lipoprotein function alterations.

**Hypothesis:** We hypothesized that FH children had disrupted lipoprotein function.

**Methods:** We analyzed LDL aggregation susceptibility and HDL-apoA-I exchange to apoA-I ratio (HAE/apoA-I ratio), and activity of four proteins that regulate lipoprotein metabolism (CETP, LCAT, PLTP and PON1) in plasma samples derived from children with FH (n = 47) and from healthy children (n = 56). Potential biological mechanisms behind any variation in lipoprotein functionalities were explored using an NMR-based metabolomics profiling approach.

**Results:** LDL aggregation was higher and HAE/apoA-I ratio was lower in FH children than in healthy children. LDL aggregation associated positively with LDL-C and negatively with triglycerides, and HAE/apoA-I ratio associated negatively with LDL-C. Generally, the metabolomic profile for LDL aggregation was a mirror image of that for HAE/apoA-I ratio.

**Conclusions:** FH children displayed increased atherogenicity of LDL and disrupted HDL function. These newly observed functional alterations in LDL and HDL may increase the risk for atherosclerotic cardiovascular disease in FH children.

## Introduction

Besides the plasma concentration, lipoprotein particle function is emerging as an important factor in the progression of atherosclerotic cardiovascular disease (ASCVD) (1,2). Low-density lipoprotein (LDL) particles undergo significant modifications *in vivo*, particularly in the unique microenvironment in the arterial wall, via oxidizing agents, proteases, and lipases (2). Modified LDL particles are prone to aggregation, and the tendency for LDL particles to aggregate is one of several metrics of LDL function, predictive of ASCVD risk (3).

Furthermore, the spectrum of high-density lipoprotein (HDL) particles and apolipoprotein A-I (apoA-I) have specific roles in reverse cholesterol transport (RCT), a process that conveys cholesterol and lipids from peripheral tissues to the liver. HDL function can be quantified by metrics of RCT, including cholesterol efflux capacity (CEC) or HDL-apoA-I exchange to apoA-I ratio (HAE/apoA-I ratio) (4), measures also predictive of ASCVD risk (5,6).

Because dyslipidemia drives ASCVD, a major cause of disability and death worldwide (1), and because lipoprotein modification connects lipids and inflammation in atherosclerosis (2), biomarkers of lipoprotein function have the potential to identify ASCVD risk well in advance of acute pathological manifestation. However, it is unclear whether children display measurable alterations in lipoprotein function. We reasoned that children with familial hypercholesterolemia (FH) would be a suitable group to examine this research question. Patients with FH show elevated plasma LDL cholesterol (LDL-C) since birth, and a variety of markers of atherosclerotic development are observable even in early childhood (7). We therefore hypothesized that FH children would have disrupted lipoprotein function.

## Subjects and methods

The present study is a cross-sectional analysis of FH children and healthy children. We have reported research findings from the same populations previously (8,9) (**Figure S1, Table S1**).

### Study design, setting and participants

Briefly, we recruited 47 FH children from the outpatient Lipid Clinic at Oslo University Hospital (OUH) in Oslo, Norway (9). All children had a definite FH diagnosis, verified by genetic or clinical diagnosis, the latter based on the Simon Broome criteria (10). Eighteen children (38 %) were currently on statins, and 20 (43 %) had LDL receptor (LDLR) negative mutations (**Supplementary Material**) (9,11). Furthermore, we included 56 non-FH, healthy children that were part of the Stork children follow-up study at Department of Nutrition, University of Oslo (UoO) in Oslo, Norway (8,12). Data collection occurred in the period September 2013 to October 2015.

All children, and their parents when the child was under 16 years, gave written informed consent to participate in the study. The Regional Committee for Research Ethics in South East Norway approved the study, and the study protocol was in accordance with the declaration of Helsinki.

### Data measurements and variables

We obtained clinical and biochemical data as previously described (8,12). Briefly, we assessed clinical variables at the time of visit, including weight, height, and statin use. We collected blood samples at time of visit, and consecutively analyzed standard biochemistry in heparin-plasma at the Department of Medical Biochemistry, OUH. Data collection for the two groups of children was conducted by different researchers but followed the same standardized research protocol to reduce the impact of information bias. Apart from during the data collection phase, all laboratory analyses (described in the following) were performed with the analysts being blinded to FH status, statin use, sex, or any other characteristics.

#### LDL aggregation assay

We analyzed LDL aggregation as previously described, given in brief in Supplementary Material (3,13). In accordance with previous reports, we herein analyzed LDL aggregation values for the two-hour timepoint. Because we normalized input to the aggregation assay to 0.2 mg apoB-100/mL, effectively adjusting for LDL-C, this represents a *per particle* (PP) metric of LDL aggregation. The LDL aggregation (PP) represents the LDL aggregation variable reported in other studies (3,13). We engineered an additional variable by multiplying LDL aggregation values by laboratory-measured LDL protein concentration. Because this second variable considers the *totality* of LDL exposure, that is, unadjusted for LDL-C, it represents a *total load* (TL) metric of LDL aggregation. Although we present both variables in figures, we consider the latter clinically most important and appropriate; hence, we mostly describe *LDL aggregation (TL)* throughout the manuscript text.

#### HDL-apoA-I exchange (HAE) assay

We analyzed the HAE/apoA-I ratio as previously described, and summarized in Supplementary Material (4). HAE/apoA-I is a measure of HDLs ability to “exchange” apoA-I, normalized relative to apoA-I plasma level. ApoA-I exchange in the intima is required for HDL biogenesis so HAE/apoA-I therefore *indirectly* reflects HDL’s ability to initiate RCT. In contrast, the cell-based CEC assay, which measures the potential of macrophages to deliver cholesterol to lipid-poor apoA-I, is considered the gold standard of HDL function assessment (14). While well established, the CEC method has not been standardized and is subject to the variability induced by cell culture methods. Despite this, the HAE/apoA-I ratio strongly correlates with CEC, and s a result the HAE/apoA-I ratio is an effective and highly reproducible surrogate biomarker of CEC (15).

#### Lipoprotein metabolism-regulating proteins

We analyzed the activity of four proteins that participate in the regulation of lipoprotein metabolism: cholesteryl ester transfer protein (CETP), lecithin–cholesterol acyltransferase (LCAT), phospholipid transfer protein (PLTP) and paraoxonase-1 (PON1). CETP activity (nmol/mL/h) was analyzed as the transfer/exchange of radiolabeled [^14^C]-cholesteryl oleate between exogenously added human LDL and HDL2, as described (16). LCAT activity (nmol/ml/h) was assessed by measuring cholesterol esterification activity using exogenous [^3^H]-cholesterol-labelled HDL proteoliposome discs as the substrate (17). PLTP activity (nmol/ml/h) was determined with a radiometric method as described (18). PON1 (umol/min) activity was measured with a chromogenic method (19).

#### Quantitative NMR metabolomics

We measured a broad set of biomarkers involved in human metabolism-related health and disease using a commercially available, high-throughput nuclear magnetic resonance (NMR) spectroscopy platform (Nightingale Health, Finland) (20). The method is thoroughly described in separate reports (20,21). Briefly, the biomarkers covered particle concentration and lipid content of 14 subclasses of lipoproteins, and plasma fatty acids, amino acids, glucose metabolites, ketone bodies and other biomarkers, including certain protein biomarkers. In contrast to our previous work which was based on the original 2016 algorithm (9), the data values for the present analysis were estimated using the 2020 algorithm (Nightingale Health, Finland). Note also that previous metabolomics analyses regarding LDL aggregation was based on LC-MS, not NMR (3).

### Data analyses

We performed all data analyses in R version 4.0.0 (22) using RStudio (Boston, MA, USA, www.rstudio.com) and the *tidyverse* framework, including data cleaning, data manipulation, data modeling, and data visualization (23). Thorough exploratory data analysis guided all data analysis decision-making described below. We refer to R packages and functions where appropriate using the following notation: `package::function`.

#### Feature engineering

We calculated body mass index (BMI) z score based on World Health Organization (WHO) Growth References (`addWGSR::zscorer`) (24). Furthermore, we addressed missing data with the following strategies. First, for CRP, estradiol, and testosterone, 80, 55 and 44 participants had values below or exactly at detection limits of 0.6 mg/L, 0.04 nmol/L and 0.4 nmol/L, respectively. We imputed these entries manually by generating a random uniform between zero and specific detection limit (`stats::runif`). Additionally, a single participant had missing entries for estradiol and testosterone. Second, for HAE/apoA-I ratio, we changed a single entry to missing because it was unlikely high (> 58), totaling 13 missing entries for FH children for this variable. Additionally, the concentration and lipid content of the largest lipoprotein particles was zero for many subjects, likely because of values being below the detection limit. For XXL-VLDL particles and lipids, 27 entries were zero (covering both FH children and healthy children); similarly, for XL-VLDL particles and lipids, entries were zero for two FH children. We changed all these to missing. Finally, we imputed missing entries with the k-nearest neighbors (kNN) algorithm within the *tidymodels* framework (`recipes::recipe` and `recipes::step_knnimpute` to impute, and `recipes::prep` and `recipes::juice` to collect the imputed dataset).

Furthermore, also prior to modeling, we loge transformed (`base::log`) all right skewed continuous exposures and outcomes to normal distributions. Right skewness was objectively defined as skewness > 1 (`e1071::skewness`). For the lipoprotein function metrics and lipoprotein metabolism-regulating proteins, this applied to the LDL aggregation variables (PP and TL) and PON1 only (see **Table S2** for the full overview); following transformations, these biomarkers displayed normal distributions except PON1, which showed a bivariate distribution, likely related to genetic variation (25). Next, we normalized all continuous variables to standard normal distributions, that is, with mean equal to zero and standard deviation equal to one (`base::scale`), to aid comparison of magnitudes and to aid visualization of the results. Consequently, the β regression coefficients for continuous variables can be interpreted as *per 1 standard deviation increase*.

#### Linear models

We examined lipoprotein function metrics and lipoprotein metabolism-regulating proteins in FH children and healthy children; however, our statistical analyses were divided into two parts, described below. We used ordinary linear regression models (`stats::lm`) to explore crude models and models adjusted for one or more of the following covariates: age, sex, BMI z score, triglycerides, statin use and mutation type. All associations we present herein are multivariable linear regression models adjusted for age, sex, and BMI z score; other univariate or multivariate models with different adjustment levels were relatively similar.

We first compared lipoprotein function metrics and lipoprotein metabolism-regulating proteins (seven outcomes) in FH children with healthy children (one exposure), for example like so: *LDL aggregation (TL) ∼ FH status + covariates*. In this set of models, we also examined FH subgroups: statin users, non-statin users, LDLR negative mutations, and other mutations. Secondly, we performed a more comprehensive analysis: we examined the variation in LDL aggregation (PP), LDL aggregation (TL) and HAE/apoA-I ratio (three outcomes) across *all* clinical variables and biomarkers covered by the NMR metabolomics platform (98-99 exposures), for example like so: *LDL aggregation (TL) ∼ L-LDL-P + covariates*. In the second part, we performed the analyses for all children combined (306 models), and separately for FH children (312 models) and healthy children (306 models), yielding a total of 924 models.

### Significance and power

For the first analysis part, we set alpha level to 0.05. In the second part, we did not consider significance in the usual way. Instead, we interpreted the direction and strength of the β regression estimates, the uncertainty around those estimates, and the relationship between variables. Still, we reported significance by standard cutoffs in the figures: P < 0.001, P < 0.01, P < 0.05 and P ≥ 0.05, to aid translation of the results.

We did not perform *a priori* power calculations for the present study. However, to give an indication of the power of our analyses, we performed simple *post hoc* power calculations for the general linear model (`pwr::pwr.f2.test`), yielding the following result. Given six degrees of freedom, for an association quantified by an R^2^ (explained variance) of 0.13 (which corresponds to the *median* R^2^ among our associations), to have 80 % power, we would have needed a *total* sample size of approximately 98, 135 and 184 subjects, for P value thresholds of 0.05, 0.01 and 0.001, respectively.

#### Miscellaneous

For the descriptive Tables S1 and **S3**, we calculated relevant summary statistics, including mean and standard deviation (for normally distributed, continuous variables), median and interquartile range (for skewed, continuous variables), and frequency and percentage (for categorical variables). We also compared groups using Independent Samples T tests or Chi-squared tests, as appropriate.

## Results

### FH children had alterations in lipoprotein function metrics, which associated with clinical parameters

We measured LDL aggregation, HAE/apoA-I ratio, and various plasma proteins that are important *in vivo* regulators of lipoprotein metabolism. Interestingly, FH children displayed higher LDL aggregation (PP), which was further enhanced when the total load of aggregating LDL (TL) was considered, and lower HAE/apoA-I ratio (**Figure 1**, Table S3). While adjusting for statin use and mutation type did not influence these associations (data not shown), non-statin users and FH children with LDLR negative mutations generally displayed a more pronounced effect (**Figure S2** and **S3**).

**Figure 1.**
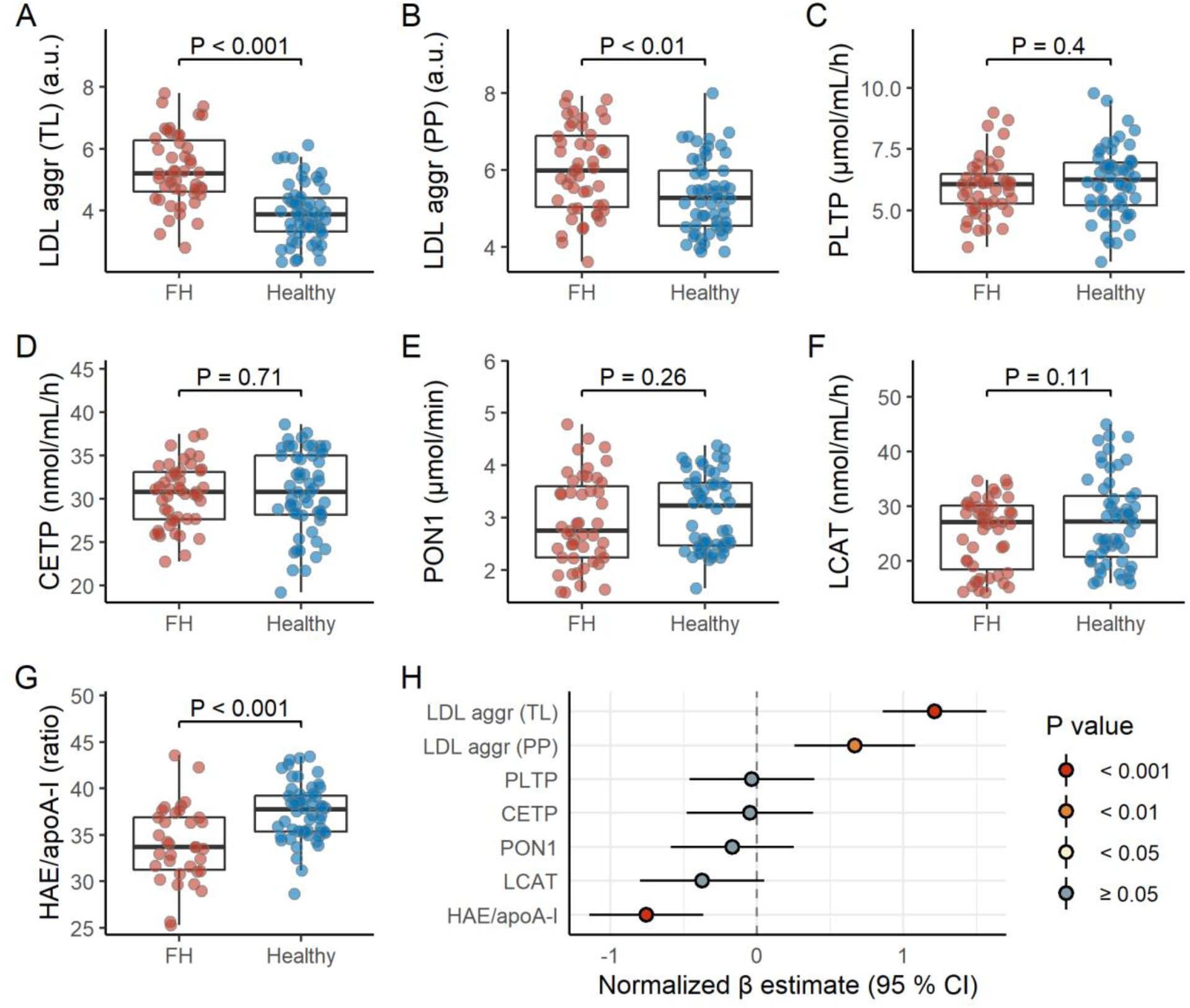
*FH children display higher LDL aggregation and lower HAE/apoA-I ratio*. Panels A-G shows raw data distributions for LDL aggregation (total load or per particle), PLTP, CEPT, PON1, LCAT, and HAE/apoA-I for FH children (n = 47) and healthy children (n = 56). Note that the LDL aggregation (total load and per particle) and PON1 variables are loge transformed. P values are based on an Independent Samples T test. See Table S3 for the summary statistics corresponding to these biomarkers for FH children and healthy children separate, and both groups combined. The forest plot (panel H) shows the estimated difference between FH children and healthy children, represented by β regression coefficients (± 95 % confidence intervals). Coefficients on the right side of the zero-line indicate higher level in FH children, and opposite for the left side. All biomarkers were normalized prior to modeling (mean = 0, standard deviation = 1); the models are adjusted for age, sex, and BMI z score. Abbreviations: aggr, aggregation; apoA-I, apolipoprotein A-I; CETP, cholesteryl ester transfer protein; HAE, HDL-apoA-I exchange; HDL, high-density lipoprotein; LCAT, lecithin–cholesterol acyltransferase; LDL, low-density lipoprotein; PLTP, phospholipid transfer protein; PON1, paraoxonase-1; PP, per particle; TL, total load.

LDL aggregation (TL) and HAE/apoA-I ratio associated with several clinical parameters (**Figure** S3 and **S4**). Specifically, LDL aggregation (TL) associated positively with LDL-C and sex hormones, and negatively with triglycerides. HAE/apoA-I ratio associated negatively with LDL-C and sex hormones, in addition to age (Figure S3 and S4). Interestingly, in general, the associations for LDL aggregation (TL) and HAE/apoA-I ratio went in the opposite directions, except for triglycerides. Of note, while the associations for triglycerides were robust in subgroup analyses, the associations for LDL-C were not (Figure S4), likely because of the limited range in LDL-C (**Figure S5**).

The lipoprotein metabolism-regulating biomarkers displayed no clear differences across groups (Figure 1), although LCAT activity inversely associated with BMI and triglycerides (Figure S3).

### Lipoprotein function metrics associated with various lipid subclasses and metabolites, supporting the clinical phenotypes

Next, to explore further why LDL aggregation (PP and TL) and HAE/apoA-I ratio differed in FH children and healthy children, we investigated the association of the lipoprotein function metrics with several lipoprotein subclasses and metabolites covering key facets of human metabolism.

LDL aggregation (TL) associated positively with LDL lipoprotein subclasses, apoB, VLDL remnants (XS-VLDL and IDL) and LDL particle diameter, and inversely with major VLDL particles and the smallest HDL particles (**Figure 2**, Table S2). Like for the clinical parameters (Figure S3), LDL aggregation (TL) and HAE/apoA-I ratio patterns were largely mirror images of each other. Furthermore, cholesterol fractions in LDL and VLDL (total, esterified or free) associated positively with LDL aggregation (TL), and so did phospholipids in LDL, plasma total sphingomyelins (SM), and the SM/phosphatidylcholine (PC) ratio (**Figure 3**, Table S2). The proportion of triglycerides to total lipids (LDL-TG %) and the ratio of CE to FC in LDL particles (LDL-CE/FC) negatively associated with LDL aggregation (TL). In contrast, for HAE/apoA-I ratio, most of the associations described above were in the opposite direction. Additionally, triglycerides in VLDL or HDL and phospholipids in HDL associated negatively with LDL aggregation (TL). Even more detailed subclass analyses showed the same trends (data not shown).

**Figure 2.**
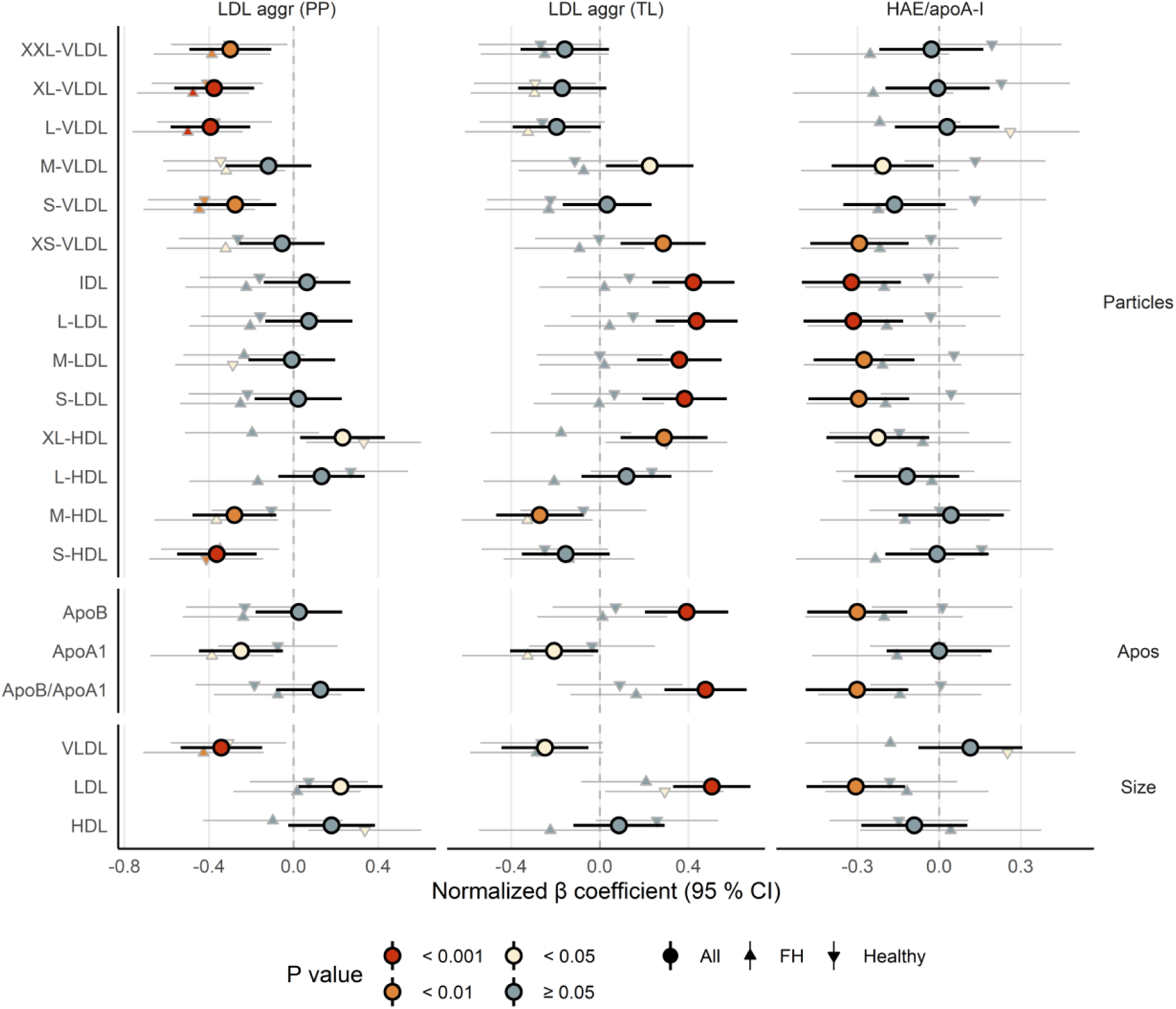
*LDL aggregation and HAE/apoA-I ratio associate with lipoprotein particles, apolipoproteins, and particle size*. The forest plot shows the association between lipoprotein function metrics (LDL aggregation (per particle or total load) or HAE/apoA-I ratio) and a subset of NMR-derived metrics (lipoprotein particles, apolipoproteins and lipoprotein particle size, for the major lipoprotein subclasses), represented by β regression coefficients (± 95 % confidence intervals). The symbols show the associations for all children combined (larger circles), FH children only (upward pointing triangles), and healthy children only (downward pointing triangles). Coefficients on the right side of the zero-line indicate a positive association, and opposite for the left side. Lipoprotein function metrics and all NMR metrics were normalized prior to modeling (mean = 0, standard deviation = 1); the models were adjusted for age, sex, and BMI z score. Abbreviations: aggr, aggregation; apoA-I, apolipoprotein A-I; ApoB, apolipoprotein B; HAE, HDL-apoA-I exchange; HDL, high-density lipoprotein; IDL, intermediate-density lipoprotein; L, large; LDL, low-density lipoprotein; M, medium; PP, per particle; S, small; TL, total load; VLDL, very low-density lipoprotein; XL, very large; XS, very small; XXL, extremely large.

**Figure 3.**
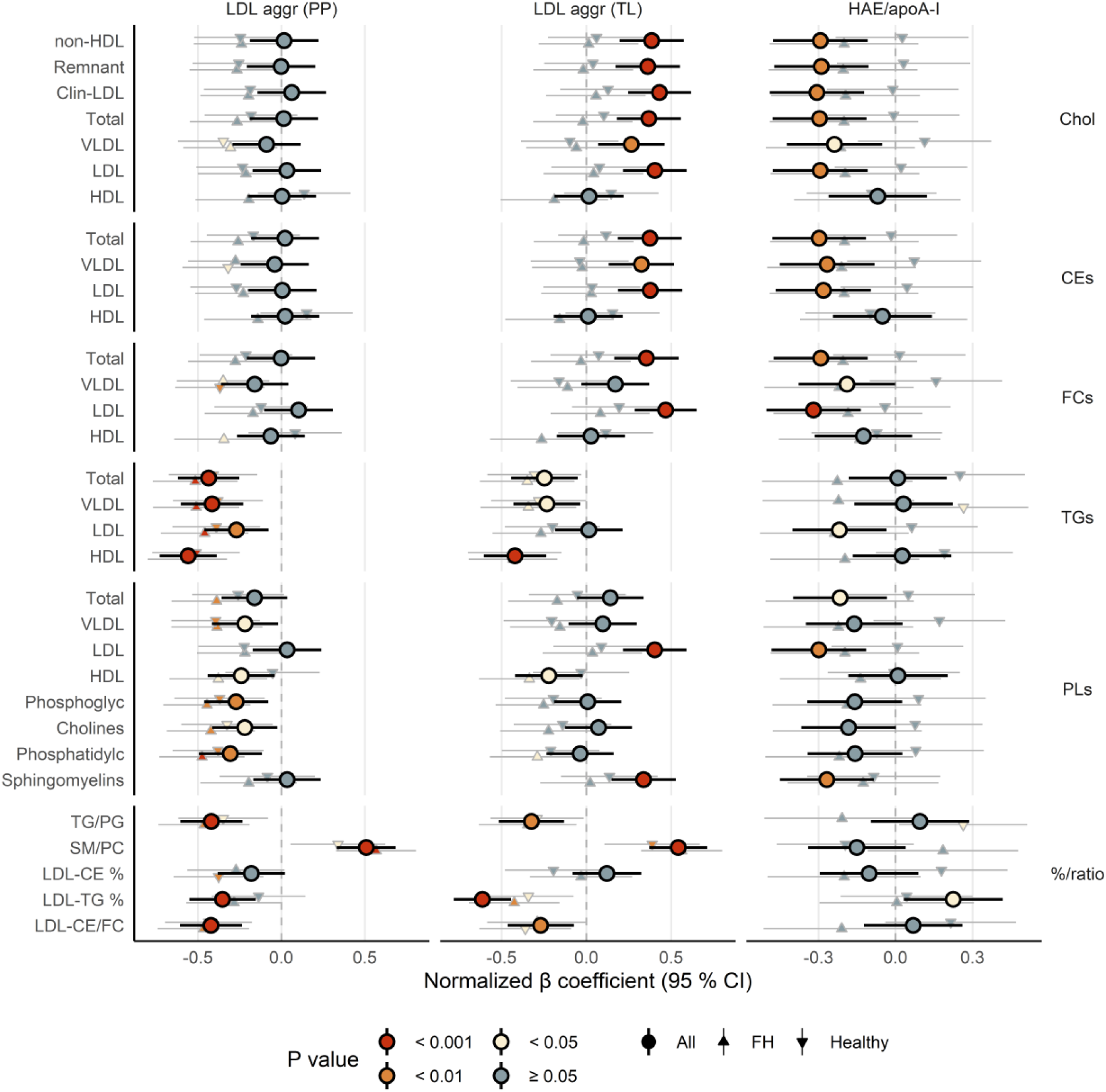
*LDL aggregation and HAE/apoA-I ratio associate with total cholesterol, cholesterol esters, free cholesterol, triglycerides, and phospholipids*. The forest plot shows the association between lipoprotein function metrics (LDL aggregation (per particle or total load) or HAE/apoA-I ratio) and a subset of NMR-derived metrics (total cholesterol, cholesterol esters, free cholesterol, triglycerides, and phospholipids, for the major lipoprotein subclasses). Interpretation similar as for Figure 2. Abbreviations: aggr, aggregation; apoA-I, apolipoprotein A-I; ApoB, apolipoprotein B; CEs, cholesterol esters; Chol, cholesterol; Clin, clinical; FCs, free cholesterol; HAE, HDL-apoA-I exchange; HDL, high-density lipoprotein; IDL, intermediate-density lipoprotein; LDL, low-density lipoprotein; Phosphatidylc, phosphatidylcholines; Phosphoglyc, total phosphoglycerides; PLs, phospholipids; PP, per particle; SM/PC, sphingomyelin/phosphatidylcholine ratio; TGs, triglycerides; TG/PG, triglyceride/phophoglycerides ratio; TL, total load; VLDL, very low-density lipoprotein.

PUFA, omega-6 and linoleic acid (LA) levels, as well as degree of unsaturation, all associated positively with LDL aggregation (TL) and negatively with HAE/apoA-I ratio (**Figure 4**, Table S2). Additionally, relative content of MUFA and SFA levels associated inversely with LDL aggregation (TL). Omega-3 markers associated neither with LDL aggregation (TL) nor with HAE/apoA-I ratio. Moreover, LDL aggregation (TL) associated inversely with levels of branched-chain amino acids (BCAAs), and positively with levels of ketone bodies (**Figure 5**, Table S2). Although HAE/apoA-I ratio again displayed associations in the opposite direction, these were less pronounced.

**Figure 4.**
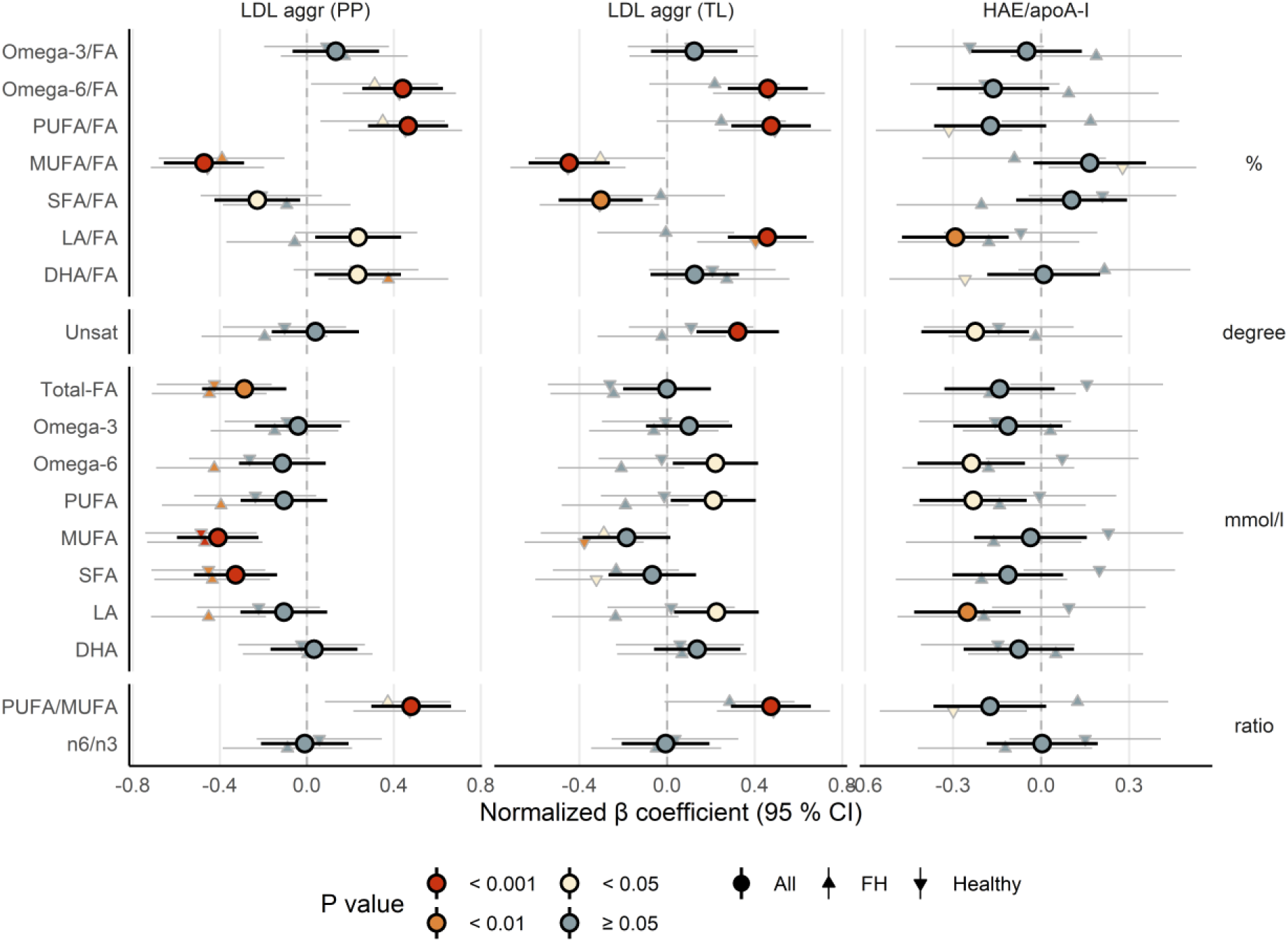
*LDL aggregation and HAE/apoA-I ratio associate with fatty acids*. The forest plot shows the association between lipoprotein function metrics (LDL aggregation (per particle or total load) or HAE/apoA-I ratio) and a subset of NMR-derived metrics (fatty acids, in absolute or relative amounts). Interpretation similar as for Figure 2. Abbreviations: aggr, aggregation; apoA-I, apolipoprotein A-I; DHA, docosahexaenoic acid; FA, fatty acids; HAE, HDL-apoA-I exchange; HDL, high-density lipoprotein; LA, linoleic acid; LDL, low-density lipoprotein; mmol/L, absolute concentration; MUFA, monounsaturated fatty acids; n6, omega-6 fatty acids; n3, omega-3 fatty acids; PP, per particle; PUFA, polyunsaturated fatty acids; SFA, saturated fatty acids; TL, total load; %, relative level (percentage of total fatty acids).

**Figure 5.**
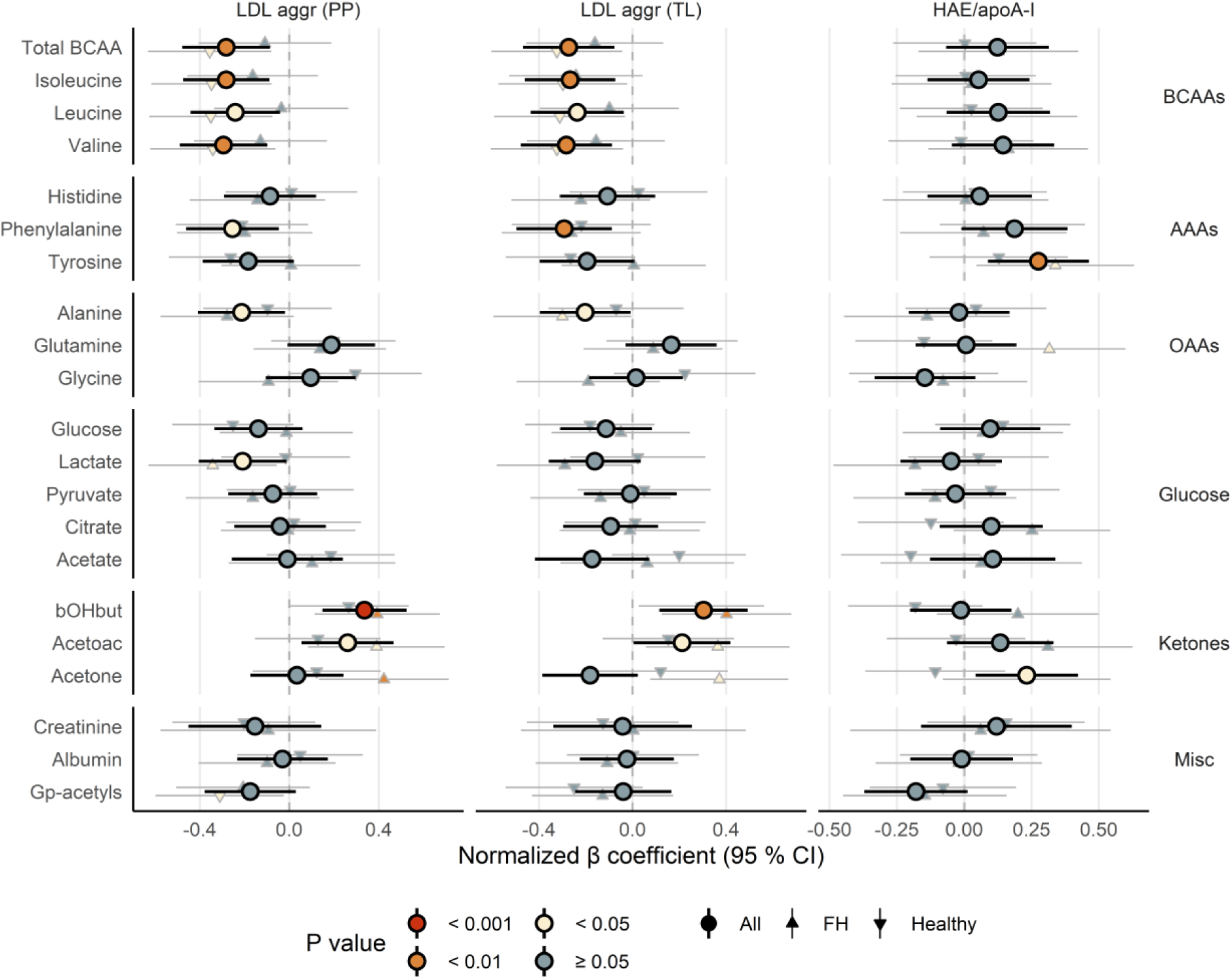
*LDL aggregation and HAE/apoA-I ratio associate with amino acids, glucose metabolites, ketone bodies and other biomarkers*. The forest plot shows the association between lipoprotein function metrics (LDL aggregation (per particle or total load) or HAE/apoA-I ratio) and a subset of NMR-derived metrics (amino acids, glucose metabolites, ketone bodies and other biomarkers). Interpretation similar as for Figure 2. Abbreviations: AAAs, aromatic amino acids; aggr, aggregation; apoA-I, apolipoprotein A-I; arom, aromatic; BCAAs, branched-chain amino acids; bOHbut, beta-hydroxybutyric acid; cap, capacity; Gp-acetyls, glycoprotein acetyls; HAE, HDL-apoA-I exchange; HDL, high-density lipoprotein; LDL, low-density lipoprotein; OAAs, other amino acids.

### Overall, biomarker associations were opposite for LDL aggregation (TL) and HAE/apoA-I ratio

With few exceptions, the associations between the lipoprotein function metrics and the lipoprotein subclasses and metabolites headed in *completely opposite* directions for LDL aggregation (TL) and HAE/apoA-I ratio (**Figure 6**, Table S2).

**Figure 6.**
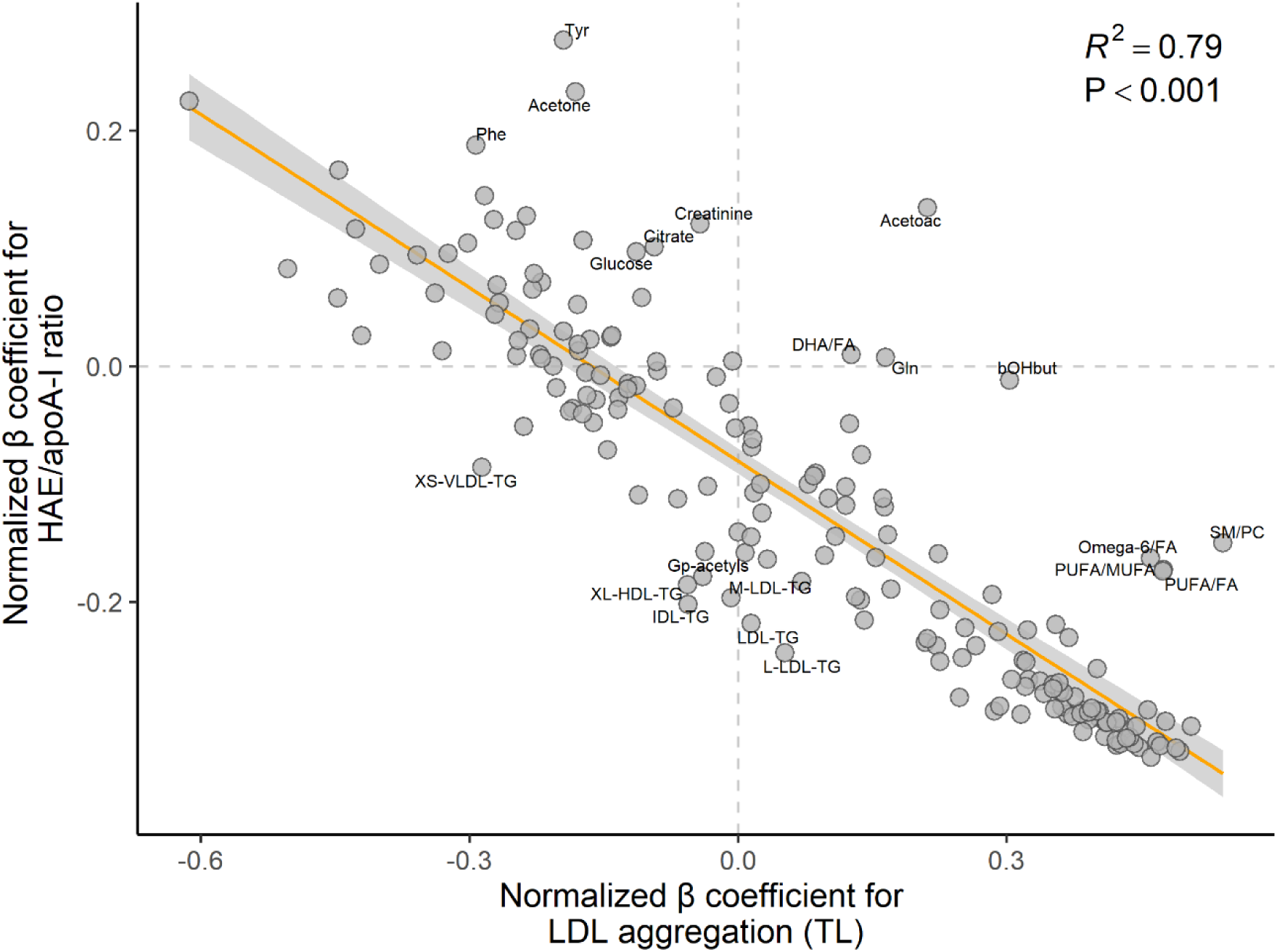
*Associations between lipoprotein function metrics and NMR-derived metrics generally go in the opposite direction for LDL aggregation and HAE/apoA-I ratio*. The scatterplot shows the inverse relationship between β regression coefficients for *all* NMR-derived metrics, for associations with LDL aggregation (TL) (x-axis) and associations with HAE/apoA-I ratio (y-axis). Note that each circle corresponds to a variable shown in Figures 2 to 5 (models for *all* children). Coefficients on the right side of the *vertical* zero-line indicate a positive association with LDL aggregation (TL), and opposite for the left side; similarly, coefficients above the *horizontal* zero-line indicate a positive association with HAE/apoA-I ratio, and opposite below. Annotated variables are those with standardized residuals above 1.5 (absolute value), which means they are more than 1.5 standard deviations away from the orange least-squares regression line. Lipoprotein function metrics and all NMR metrics were normalized prior to modeling (mean = 0, standard deviation = 1); the models were adjusted for age, sex, and BMI z score. Abbreviations: Acetoac, acetoacetate; apoA-I, apolipoprotein A-I; bOHbut, beta-hydroxybutyric acid; DHA, docosahexaenoic acid; FA, fatty acids; Gln, glutamine; Gp-acetyls, glycoprotein acetyls; HAE, HDL-apoA-I exchange; HDL, high-density lipoprotein; IDL, intermediate-density lipoprotein; L, large; LDL, low-density lipoprotein; M, medium; MUFA, monounsaturated fatty acids; P, P value; Phe, phenylalanine; PUFA, polyunsaturated fatty acids; R^2^, explained variance; TG, triglycerides; Tyr, tyrosine; VLDL, very low-density lipoprotein; XL, very large; XS, very small.

## Discussion

In the present study, we demonstrated that FH children displayed disrupted lipoprotein function compared with healthy children. Overall, the FH children were characterized by enhanced LDL aggregation (both PP and TL) and attenuated HAE/apoA-I ratio, strongly suggesting the presence of LDL particles with an increased atherogenicity and an impaired function of HDL particles in the first step of RCT, respectively. Our results indicate that variation in LDLR function, plasma LDL-C and cumulative cholesterol burden may jointly be the overarching drivers and common denominators biologically linking these metrics.

FH children had LDL particles that were particularly prone to aggregation, as reflected by high LDL aggregation (PP) in this group. Also considering the LDL protein concentration (LDL aggregation [TL]), the difference between FH children and healthy children was even more pronounced. These effects could contribute to the high risk of ASCVD in this population. Aggregation of LDL particles typically occurs following their retention in the arterial subendothelial intimal layer (2,26–28), but our present results suggest that lipoproteins in FH subjects could be primed for aggregation already while circulating in the blood, potentially due to their increased residence time in plasma caused by the underlying LDLR defect. In line with this, we previously showed that FH children have higher circulating levels of oxidized LDL (29) and display a shift in blood monocytes towards a more pro-inflammatory phenotype (12). We also observed previously that peripheral blood mononuclear cells (PBMCs) in FH children are characterized by a pro-inflammatory phenotype, which can be partially normalized upon initiation of statin treatment (30). Along with the isolated hypercholesterolemia, these alterations in FH subjects could help explain their elevated risk of ASCVD (31–33). Indeed, in the Finnish Corogene Study, baseline LDL aggregation (PP) was higher in adult ASCVD patients who died during a 2.5-year follow-up period, compared to those with stable coronary artery disease (3). In comparison, both of these patient groups had higher LDL aggregation (PP) than healthy subjects in the Health 2000 Study (3).

FH children had lower HAE/apoA-I ratio compared with healthy children, which suggests that hypercholesterolemia impairs not only LDL function, but also one of the major HDL-associated atheroprotective functions, namely, cholesterol efflux from macrophages to HDL (34). These results support our previous data (12,35). Importantly, in adult non-FH subjects CEC associated with atherosclerosis and predicted future ASCVD events independent of plasma HDL-C concentration (5,6,36). In those analyses, the researchers adjusted for several covariates including LDL-C and concluded that the association with ASCVD was independent of classical risk factors. The present results suggest that prolonged exposure to high LDL-C in otherwise healthy children has an adverse effect on HDL function, HDL metabolism, and RCT. One mechanism could be related to oxidized LDL (34,37). Work in experimental animals interestingly suggests that lack of functional hepatic LDLRs attenuate RCT via concerted action of the HDL-LDL-axis (38). Regardless of the mechanism, findings in adolescent FH subjects support that HDL function is altered early in life (39). The risk factor-related detrimental effect on RCT is likely not isolated to high LDL-C, though, as we previously observed similar alterations upon prolonged exposure to hyperglycemia in children with diabetes mellitus type I (4).

Clinical and biomarker associations for LDL aggregation (TL) and HAE/apoA-I ratio were largely mirror images of each other, suggesting that these features correlate. In our study populations, this was likely mediated by the variation in the concentration of circulating LDL particles and associated downstream effectors such as oxidized LDL. Notably, for LDL aggregation (TL), the strength and direction of associations and interrelations across all variables were practically identical to those seen for isolated hypercholesterolemia (9); for HAE/apoA-I ratio, associations were in the opposite direction. This included LDL lipoprotein particle subclasses, apoB, LDL diameter, cholesterol fractions in LDL and VLDL (total, esterified, free), phospholipids in LDL, SMs, PUFA, omega-6, and LA. Furthermore, although both LDL aggregation (TL) and HAE/apoA-I ratio associated with LDL-C, we observed an unexpected relationship with FH status: the results were *not* robust in analyses of FH children and healthy children separately (Figures 2-5 and S4). This Simpson’s paradox-like behavior could, however, be explained by two factors: that subgroup analyses in this case would equate to adjusting for the mediator in the causal pathway (that is, LDL-C), and the limited range in LDL-C observed *within* each group (Figure S5). In contrast, the effects of the mutation type and statin use on the studied metrics (LDL aggregation [TL] and HAE/apoA-I ratio) were in the expected directions: when compared with healthy children, LDLR negative mutations and lack of statin use were more detrimental, and other LDLR mutations and statin use were less detrimental. Taken together, it seems likely that variation in LDLR function, plasma LDL-C and cumulative cholesterol burden may jointly be the overarching drivers and common denominators biologically linking these metrics. Consequently, these findings could be extrapolated to variation in cholesterol burden in non-FH populations.

Paradoxically, LDL aggregation associated *inversely* with triglycerides, suggesting that higher triglyceride levels within the normal triglyceride range were protective. The Spearman’s correlation coefficients for LDL aggregation and triglycerides were -0.39 (PP) and -0.25 (TL), respectively. This finding was consistent in subgroup analyses, and is also corroborated by similar results from other studies: in the Health 2000 Study and the Corogene Study, the correlations between LDL aggregation (PP) and triglycerides were -0.28 and -0.38, respectively (3). The in-depth exploration of biological mechanisms for this association (Figures 2-5) only reinforced these clinical findings (Figures S3 and S4): the overall metabolomics pattern corresponded well with the expected changes paralleling lower triglycerides. For example, low triglyceride levels typically associate with low levels of triglycerides in VLDL, IDL, LDL and HDL particles, VLDL lipoprotein particle subclasses, VLDL diameter, phospholipids and cholesterol fractions in VLDL (total, esterified, free), total phosphoglycerides, total cholines, PCs, SMs, and the SM/PC ratio. Low triglycerides also associate with low SFA and MUFA and higher ketone bodies, likely corresponding to higher hepatic beta-oxidation and ketogenesis and lower liver fat content (40). Finally, low triglycerides associate with low levels of branched-chain amino acids, probably related to higher muscle beta-oxidation (41). All these observations are likely downstream effects of high insulin sensitivity which parallel low triglycerides (40). Taken together, triglycerides strongly, consistently, and *inversely* associated with LDL aggregation (see Supplementary Material for further discussion).

The present work has certain limitations that warrant mention. First, this is an observational, cross-sectional analysis, thus, we can neither infer causality nor rule out residual confounding factors. However, because FH is a well-characterized human genetic disorder, we feel confident that the LDLR mutations caused an increase in LDL-C prior to all other alterations (42). Also, the effect of confounding factors is likely lower in children than in adults, regardless of FH status. Second, the number of study participants was low, which increases the probability of false positive and negative findings. To meet this issue, we did not emphasize significance, but rather focused on the direction and strength of associations, the uncertainty around the point estimates, and their interrelations. Third, the two groups of children were not collected for a single study; rather, they were part of separate recruitments, which likely introduced bias in both data collection and standard clinical and biochemical data measurements. However, we performed data collection during a single period and adhered to strict study protocols to attenuate the potential bias. Also, the lipoprotein function metrics, lipoprotein metabolism-regulating proteins and NMR metrics were analyzed collectively by highly standardized protocols. Importantly, in this phase the analysts were blinded to the subject characteristics. Finally, we did not perform LC-MS in-depth proteomic and lipidomic analyses of the LDL particles, which could have shed light both on the triglyceride paradox and whether the specific characteristics of the particles represented a shared mechanistic link between LDL aggregation and HDL efflux.

To the best of our knowledge, this is the first study that comprehensively examines LDL and HDL lipoprotein function metrics in FH children, thus expanding our knowledge and understanding about these relevant biomarkers of ASCVD risk in severe early-life hypercholesterolemia. Although FH is a genetic disorder, the atherosclerotic process is similar in all humans, which to a large degree enables translation of our findings to the general population.

In conclusion, FH children were characterized by disrupted lipoprotein function compared with healthy children, which was related to differences in plasma lipids and lipoproteins. Higher LDL-C associated with both higher LDL aggregation (TL) and lower HAE/apoA-I ratio, suggesting that LDLR function and plasma LDL-C may be the overarching drivers and common denominators linking these metrics in a biologically relevant context. For more strict causal verification, these molecular aspects need further detailed mechanism-based investigations.

## Supporting information

STROBE checklist

## Data Availability

Summary statistics are available in Supplementary Material, but individual level raw data will not be made available.

## Abbreviations

apoA-I: apolipoprotein A-I
apoB: apolipoprotein B
ASCVD: atherosclerotic cardiovascular disease
BMI: body mass index
BCAAs: branched-chain amino acids
CETP: cholesteryl ester transfer protein
DHA: docosahexaenoic acid;
FA: fatty acids
FH: familial hypercholesterolemia
HAE: HDL-apoA-I exchange
HDL: high-density lipoprotein
LA: linoleic acid
LCAT: lecithin–cholesterol acyltransferase
LDL: low-density lipoprotein
LDL aggr (PP): LDL aggregation (per particle)
LDL aggr (TL): LDL aggregation (total load)
LDLR: LDL receptor
MUFA: monounsaturated fatty acids
n3: omega-3 fatty acids
n6: omega-6 fatty acids
PC: phosphatidylcholine
PLTP: phospholipid transfer protein
PON1: paraoxonase-1
PUFA: polyunsaturated fatty acids
RCT: reverse cholesterol transport
SFA: saturated fatty acids
SM: sphingomyelins
VLDL: very low-density lipoprotein

## Acknowledgements

We would like to thank all children and families that kindly chose to participate in our research studies. We would also like to thank technician Maija Atuegwu for helpful assistance with laboratory analyses.

## Sources of support

This work was funded by the University of Oslo (Oslo, Norway), the National Advisory Unit on FH at OUH (Oslo, Norway), the Throne-Holst Foundation for Nutrition Research (Oslo, Norway), the South-Eastern Regional Health Authority (Oslo, Norway), the Academy of Finland (#315568 and #332564 to KÖ), the Jane and Aatos Erkko Foundation (to MJ), the Finnish Foundation for Cardiovascular Research (to MJ and KÖ), the Aarne Koskelo Foundation (to KÖ), and the Novo Nordisk Foundation (to KÖ). Wihuri Research Institute is maintained by the Jenny and Antti Wihuri Foundation.

## Conflicts of interest

Dr. Christensen has received research grants and/or personal fees from Mills DA, unrelated to the content of this manuscript. Drs. Ruuth, Kovanen and Öörni have applied for a patent on the LDL aggregation method; otherwise they have no other relevant financial relationships to disclose. Dr. Ulven has received research grants from Tine DA, Mills DA, and Olympic Seafood, none of which are related to the content of this manuscript. Dr. Retterstøl reports personal fees from Amgen, Mills DA, The Norwegian Medical Association, The Norwegian Directorate of Health, Sanofi, Takeda, Chiesi, Bayer, MSD, and research grants from Oslo Economics and Mills outside the submitted work. Dr. Heier has received a research grant from the Norwegian Diabetes Association for analyses included in this manuscript. Dr. Holven has received research grants and/or personal fees from Tine DA, Mills DA, Olympic Seafood, Amgen, Sanofi, and Pronova, none of which are related to the content of this manuscript. The other authors have no relevant financial relationships to disclose.

## Authorship

Conception and design: JJC, IN, SMU, KÖ, KBH; data collection: JJC, IN, MR, MH, MJ, MPB, MO, CW, KR; data analysis: JJC, IN, MR, MH, MJ, PK, MO, KÖ, KBH; data interpretation: all authors, wrote paper and responsibility for final content: JJC, KÖ, KBH; all authors read, critically revised and approved the final manuscript.

## Notes

### Author Declarations

The Regional Committee for Research Ethics in South East Norway approved the study, and the study protocol was in accordance with the declaration of Helsinki.

