## Supplementary material for "Children with familial hypercholesterolemia display changes in LDL and HDL function: a cross-sectional study": Online Supplementary Material - Text

*Shared last authorship

1 Norwegian National Advisory Unit on Familial Hypercholesterolemia, Department of Endocrinology, Morbid Obesity and Preventive Medicine, Oslo University Hospital, Forskningsveien 2B, 0373 Oslo, Norway

2 Department of Nutrition, Institute of Basic Medical Sciences, University of Oslo, Sognsvannsveien 9, 0372 Oslo, Norway

3 Atherosclerosis Research Laboratory, Wihuri Research Institute, Helsinki, Finland

4 Research Programs Unit, Faculty of Medicine, University of Helsinki, Finland

5 Pediatric Department, Oslo University Hospital Ullevaal, Oslo, Norway

6 Oslo Diabetes Research Centre, Oslo, Norway

7 Minerva Foundation Institute for Medical Research and National Institute for Health and Welfare, Biomedicum 2U, Helsinki, Finland

8 Unit for Cardiac and Cardiovascular Genetics, Oslo University Hospital, Norway

9 Research Institute of Internal Medicine, Oslo University Hospital Rikshospitalet, 0424 Oslo, Norway

10 Faculty of Medicine, University of Oslo, 0424 Oslo, Norway

11 Seer BioLogics, Inc., Fairfield, CA, 94534, United States

12 The Lipid Clinic, Oslo University Hospital, Norway

13 Molecular and Integrative Biosciences Research Programme, Faculty of Biological and Environmental Sciences, University of Helsinki, Finland

**Extended Supplementary Methods**

In this section, we provide an extension of Methods in the main manuscript.

*Study design, setting and participants*

FH children with a known mutation in the LDL receptor (LDLR) were categorized into two groups as previously described (1,2) : “LDLR negative mutations” (defined as class 1 and 2A mutations, which largely comprised nonsense, splice site mutations, or large rearrangements), and “Other mutations” (defined as class 2B, 3, 4 and 5, comprising all other LDLR defective mutations).

*Data measurements and variables*

LDL aggregation assay

We analyzed LDL aggregation as previously described, here given in brief (3,4). First, we isolated LDL particles from plasma by $D_2O$-based sequential ultracentrifugation, and determined LDL protein concentration with the BCA protein assay (Pierce, Rockford, USA), using bovine serum albumin as a standard. Samples were then diluted to give a final concentration of 0.2 mg apoB-100/mL in 20 mM MES, pH 5.5, containing 150 mM NaCl, 2mM $MgCl_2$, 2 mM $CaCl_2$, and 50 µM $ZnCl_2$, and human recombinant secretory sphingomyelinase (hrSMase), produced in-house (4) was added to give a final enzyme concentration of 75 µg/mL. The final mixture was incubated at +37 °C; LDL aggregate size was determined immediately, and then hourly up to six hours, using dynamic light scattering (Wyatt DynaPro Plate Reader II; Wyatt Technology, CA). Aggregate size time curves were constructed based on all data points for an individual subject, and aggregate sizes after incubation for 2 hours was used in further analyses.

Lipoprotein function

See main manuscript text for further details about interpretation of the LDL aggregation variables.

HDL-apoA-I exchange (HAE) assay

We analyzed relative efficiency of apoA-I exchange by HAE/apoA-I ratio as previously described, given in brief next (5). First, we precipitated apoB-containing lipoproteins by the addition of polyethylene glycol 6000 and centrifugation. We then added 3 mg/mL nitroxide spin-labeled apoA-I probe to the isolated supernatant, and incubated at 37 °C for 15 min. The sample plus nitroxide spin-labeled apoA-I probe arrived at equilibrium, and we then scanned the intensity signal by electron paramagnetic resonance (EPR) spectroscopy with a Bruker eScan EPR spectrometer (Bruker BioSpin GmbH, Karlsruhe, Germany), outfitted with a temperature controller (Noxygen Science Transfer & Diagnostics GmbH, Elzach, Germany) (scanned at 37 °C). We compared the peak amplitude of the nitroxide signal from the HAE probe in the sample (3462-3470 Gauss) to that of a proprietary internal standard (3507-3515 Gauss) provided by Bruker, which standardizes the HAE activity values (HAE sample/standard ratio). We estimated maximum %-HAE activity by comparing HAE activity to the standard curve for the probe-lipid-associated signal. We used the average of duplicates as the final HAE output values. These were then normalized for variation in apoA-I concentration in plasma.

See main manuscript text for further details about interpretation of the HAE/apoA-I variable.

**Extended Supplementary Discussion**

In this section, we provide an extension of the Discussion in the main manuscript.

*Lower HAE/apoA-I ratio in FH children compared with healthy children suggests impaired RCT; while the molecular mechanisms are unknown, they are likely multi-faceted*

The molecular mechanisms explaining why chronically elevated LDL-C in FH would cause lower HAE/apoA-I ratio is unknown but likely complex and multi-faceted. For example, under physiological conditions, the LDLR pathway is an important *parallel* route of RCT: CETP facilitates flux of cholesterol from HDL to VLDL and LDL particles, which are subsequently taken up by hepatocytes (6,7). In heterozygous FH, however, LDLR capacity is approximately 50 % of normal (at least for subjects with LDLR negative mutations), resulting in congestion and accumulation of apoB-containing particles, thereby disturbing this pathway (8,9). Variation in LDLR expression and plasma LDL-C in non-FH subjects could contribute to similar alterations, although to a lesser extent. Furthermore, variation in LDLR expression and plasma LDL-C could also affect LXR-driven expression of ABC transporters (10). In observational studies, chronic exposure to an unhealthy diet, elevated risk factors, and prevalent disease consistently associate with *reduced* expression of ABC transporters in whole blood or PBMCs (11–13). This is relevant since ABC transporters and LCAT in a concerted fashion generate mature, spherical HDL particles by first transporting cellular cholesterol from macrophages via ABCA1 to lipid-poor apoA-I and discoidal pre-β-HDL particles, followed by immediate LCAT-facilitated cholesterol esterification, entry of cholesterol esters in HDL particle core and finally HDL particle hepatic uptake. Of note, the cholesterol esterification by LCAT is affected by the lipidome of the HDL particle, particularly SM which is a physiological inhibitor of the esterification reaction (14).

*LDL aggregation and HAE/apoA-I ratio could represent biomarkers of risk factor burden, and they respond to lifestyle and pharmacological interventions*

Collectively, LDL aggregation and HAE/apoA-I ratio could represent non-causal biomarkers of atherosclerotic risk factor burden. For example, LDL aggregation (PP) improved quickly following improvement in dyslipidemia by lifestyle or pharmacological interventions, an effect likely mediated via changes in SM and PC content of the LDL particles (3). Similarly, cholesterol efflux improved following gastric bypass surgery (15) or upon initiation of a Mediterranean diet (16). We also recently showed that PBMC gene expression of ABCG1 increased very consistently paralleling a reduction in plasma LDL-C, in an 8-week intervention trial of improved dietary fatty acid quality (17). In contrast, in the short-term post-prandial setting, PBMC gene expression of LXR target genes including ABC transporters paradoxically *increased more* upon an SFA challenge than with a PUFA challenge, illustrating the importance of considering the *temporal* effects of metabolic regulation in search of such molecular biomarkers (18,19).

*Paradoxically, triglycerides inversely associated with LDL aggregation, likely because of triglyceride-driven alterations in the lipidome and proteome of the LDL particles*

It is unclear which specific mechanisms caused the inverse association between triglycerides and LDL aggregation (both PP and TL). It is well known, however, that variation in plasma triglycerides affects other lipid compartments; indeed, triglycerides associate positively with proportion of triglycerides and PC in LDL particles, and negatively with proportion of SM. The proportion of these lipids has been shown to impact LDL aggregation (3). Increased proportion of LDL-SM content increases LDL aggregation whether it is induced by oxidation, proteolysis, or lipolysis by SMase or phospholipase A2 (3). In the *ex vivo* LDL aggregation assay, hrSMase hydrolyzes SMs in the surface monolayer of LDL particles, which leads to generation of ceramides, which subsequently induces dramatic conformational changes in apoB-100, ensuing surface exposure of hydrophobic domains of the protein. The protein components of several LDL particles then physically bind to each other by hydrophobic interactions, inducing formation of very-large, micron-sized LDL aggregates (20). Such large ceramide-containing LDL aggregates are found in human atherosclerotic lesions (21). The proportions of the two major phospholipids, PC and SM, appear to control the conformation of apoB-100 on LDL particles (3), but whether the triglycerides contained in LDL particles also influence the propensity of the particles to aggregate, is currently unknown.

**Supplementary Tables**

*Supplementary Table 1*

**Table S1**. Clinical characteristics.

| | All children (n = 103)[1] | FH children (n = 47)[2] | Healthy children (n = 56) | P[3] |
|---|---|---|---|---|
| Statin, yes (%) | 18 (18) | 18 (38) | | |
| LDLR negative mutation, n (%) | 20 (19.4) | 20 (42.6) | | |
| Sex, n girls (%) | 54 (52) | 26 (55) | 28 (50) | 0.73 |
| Age, years | 11 (3) | 13 (4) | 10 (2) | ***<0.001*** |
| BMI, z score | 0.42 (0.96) | 0.44 (0.94) | 0.40 (0.98) | 0.82 |
| Total-C, mmol/L | 4.7 (1.5) | 5.4 (2) | 4.0 (0.8) | ***<0.001*** |
| LDL-C, mmol/L | 2.7 (1.5) | 3.6 (1.8) | 2.1 (0.9) | ***<0.001*** |
| HDL-C, mmol/L | 1.5 (0.3) | 1.5 (0.3) | 1.5 (0.3) | 0.25 |
| Triglycerides, mmol/L | 0.7 (0.4) | 0.8 (0.3) | 0.7 (0.4) | 0.97 |
| Glucose, mmol/L | 5.2 (0.6) | 5.1 (0.6) | 5.2 (0.5) | 0.17 |
| CRP, mg/L | 0.7 (0.5) | 0.7 (0.6) | 0.7 (0.5) | 0.37 |
| Estradiol, nmol/L | 0.28 (0.47) | 0.15 (0.20) | 0.37 (0.50) | ***<0.01*** |
| Testosterone, nmol/L | 0.5 (0.5) | 0.6 (2.1) | 0.5 (0.6) | ***<0.01*** |

Continuous variables are reported as median (IQR), except for age, BMI, HDL-C and glucose, which are mean (SD). Abbreviations: n, number of participants; BMI, body mass index; C, cholesterol; LDL, low-density lipoprotein, HDL, high-density lipoprotein; CRP, C-reactive protein.

[1]n=92 for estradiol and testosterone.

[2]n=46 for estradiol and testosterone.

[3]P values are based on an Independent Samples T test, except for sex, which is a Chi-squared test. Bold-italics indicates P < 0.05.

*Supplementary Table 2*

**Supplementary Table 2**. Explanatory models. Due to document size constraints, Supplementary Table 2 can be found in a separate Excel file only.

*Supplementary Table 3*

**Table S3**. Biomarker distributions.

| | All children (n = 103)[1] | FH children (n = 47)[2] | Healthy children (n = 56) | P[3] |
|---|---|---|---|---|
| LDL aggregation (TL), a.u. | 81 (160) | 180 (430) | 48 (54) | ***<0.001*** |
| LDL aggregation (PP), a.u. | 240 (540) | 400 (830) | 190 (310) | ***<0.01*** |
| HAE/apoA-I ratio, a.u. | 36 (4) | 34 (4) | 37 (3) | ***<0.001*** |
| CETP, nmol/mL/h | 31 (4) | 30 (4) | 31 (5) | 0.71 |
| PLTP, nmol/mL/h | 6100 (1300) | 6000 (1200) | 6200 (1400) | 0.40 |
| PON1, umol/min | 17 (28) | 16 (27) | 25 (27) | 0.69 |
| LCAT, nmol/ml/h | 26 (7) | 25 (6) | 27 (8) | 0.11 |

Continuous variables are reported as mean (SD), except for LDL aggregation and PON1, which are median (IQR). Abbreviations: a.u., arbitrary units; CETP, cholesteryl ester transfer protein; HAE/apoA-I, high-density lipoprotein-apolipoprotein A-I exchange adjusted for apolipoprotein A-I; LCAT, lecithin–cholesterol acyltransferase; LDL, low-density lipoprotein; PLTP, phospholipid transfer protein; PON1, paraoxonase-1.
[1]n=90 for HAE/apoA-I ratio.
[2]n=34 for HAE/apoA-I ratio.
[3]P values are based on a Independent Samples T test. Bold-italics indicates P < 0.05.

**Supplementary Figures**

*Supplementary Figure 1*

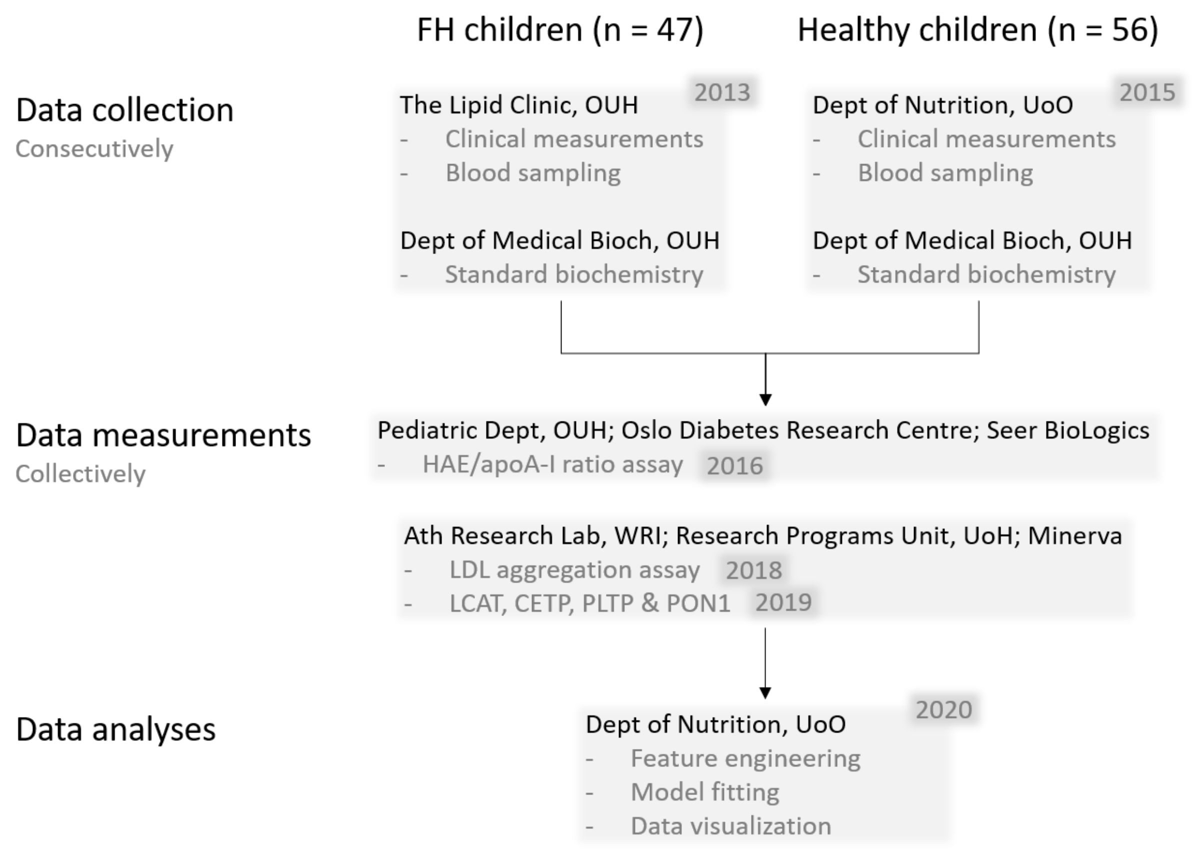


**Supplementary Figure 1**. *Participant flow chart*. The flow chart describes the different study phases: data collection including biochemical analyses, data measurements in focus in the present work, and data analyses. The figure corresponds to the descriptions in Methods. Abbreviations: ApoA-I, apolipoprotein A-I; Ath, Atherosclerosis; Bioch, Biochemistry; CETP, Cholesteryl ester transfer protein; Dept, department; FH, familial hypercholesterolemia; HAE, HDL-apoA-I exchange; HDL, high-density lipoprotein; LCAT, Lecithin–cholesterol acyltransferase; LDL, low-density lipoprotein; Minerva, Minerva Foundation Institute for Medical Research; OUH, Oslo University Hospital; PLTP, Phospholipid transfer protein; PON1,

Paraoxonase and arylesterase 1; UoH, University of Helsinki; UoO, University of Oslo; WRI, Wihuri Research Institute.

*Supplementary Figure 2*

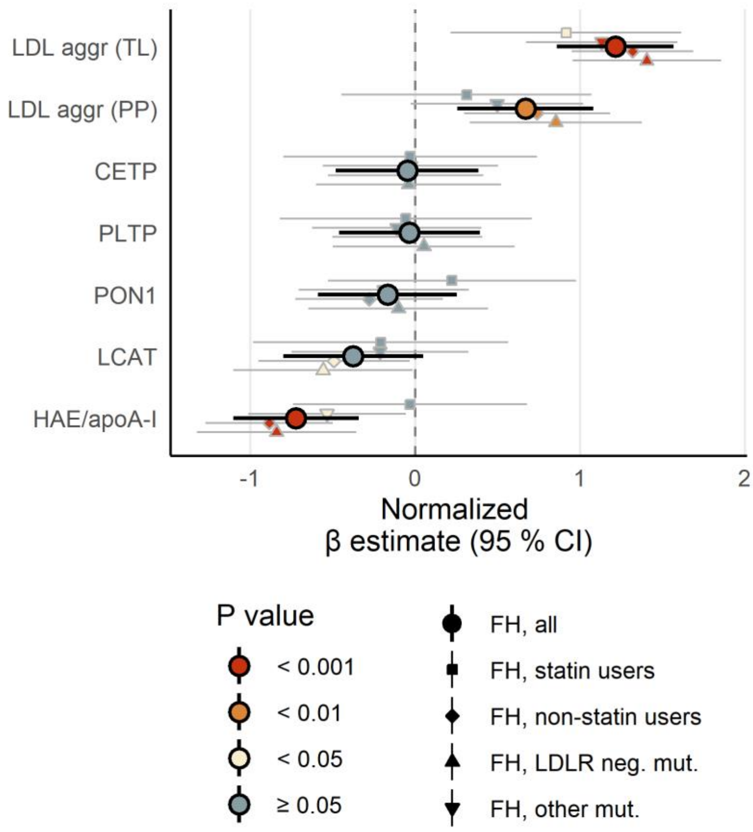


**Supplementary Figure 2.** *FH children display higher LDL aggregation and lower HAE/apoA-I ratio, although both statin use and mutation type modify these associations*. The forest plot shows the estimated difference between FH children (either all of them or subgroups of FH children) and healthy children, represented by β regression coefficients (± 95 % confidence intervals). Coefficients on the right side of the zero-line indicate higher level in FH children, and opposite for the left side. All biomarkers were normalized prior to modeling (mean = 0, standard deviation = 1); the models are adjusted for age, sex and BMI z score. Abbreviations: aggr, aggregation; apoA-I, apolipoprotein A-I; CETP, cholesteryl ester transfer protein; HAE, HDL-apoA-I exchange; HDL, high-density lipoprotein; LCAT, lecithin–cholesterol acyltransferase; LDL, low-density lipoprotein; PLTP, phospholipid transfer protein; PON1, paraoxonase-1; PP, per particle; TL, total load.

*Supplementary Figure 3*

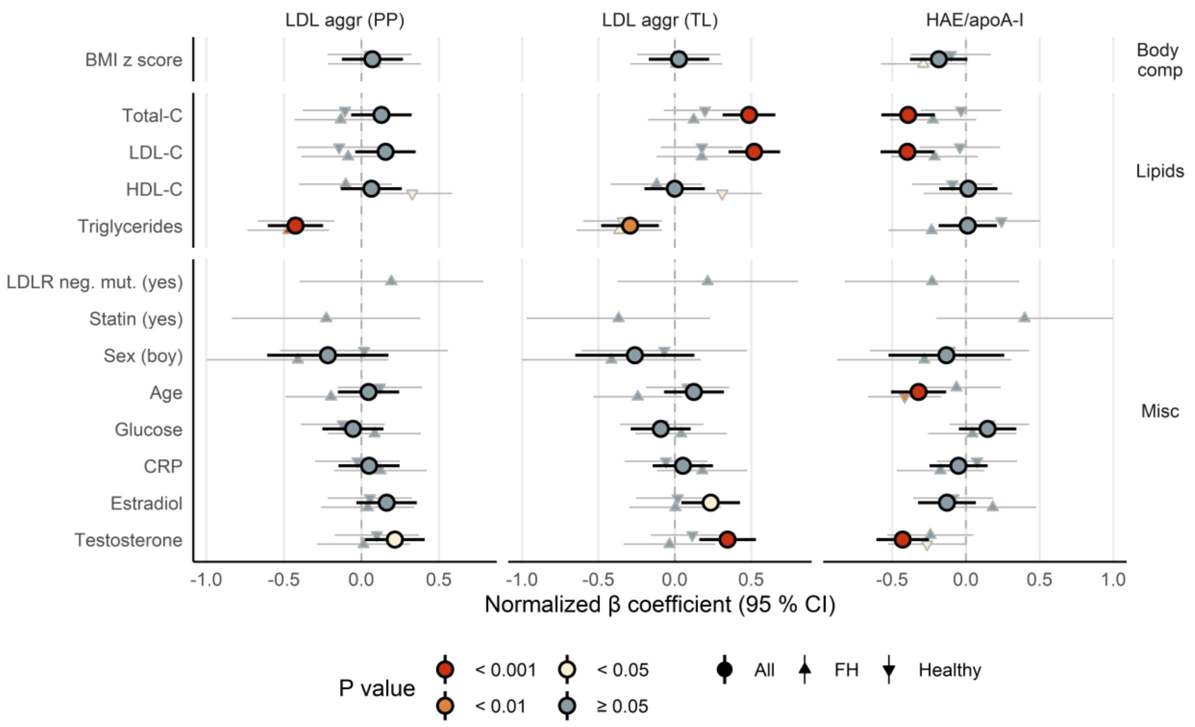


**Supplementary Figure 3**. *LDL aggregation and HAE/apoA-I ratio associate with clinical parameters*. The figure shows the association between measures of lipoprotein function (LDL aggregation (per particle or total load) or HAE/apoA-I ratio) and various clinical parameters, represented by β regression coefficients (± 95 % confidence intervals). The symbols show the associations for all children combined (larger circles), FH children only (upward pointing triangles), and healthy children only (downward pointing triangles). Coefficients on the right side of the zero-line indicate a positive association, and opposite for the left side. Measures of lipoprotein function and all clinical parameters were normalized prior to modeling (mean = 0, SD = 1); the models were unadjusted (univariable) associations. Abbreviations: aggr, aggregation; apoA-I, apolipoprotein A-I; BMI, body mass index; C, cholesterol; CRP, C-reactive protein; HAE, HDL-apoA-I exchange; HDL, high-density lipoprotein; LDL, low-density lipoprotein; LDLR, LDL receptor; mut, mutation; neg, negative; PP, per particle; TL, total load.

*Supplementary Figure 4*

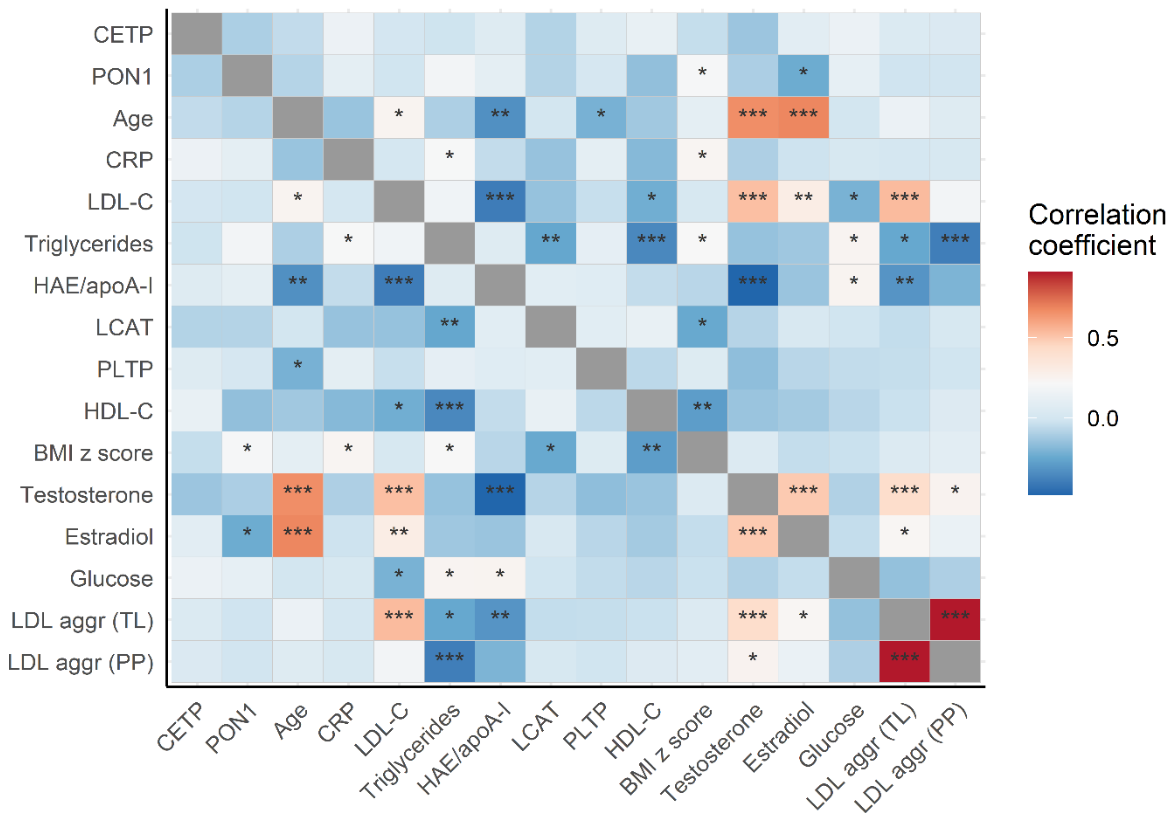


**Supplementary Figure 4**. *LDL aggregation and HAE/apoA-I associate with clinical parameters*. The figure shows a heatmap of bivariate Spearman's correlation coefficients (all children). Skewed variables were log$_e$ transformed before correlation. Stars correspond to significance level: ***, P < 0.001; **, P < 0.01, *, P < 0.05. The rows and columns have been ordered using a clustering algorithm, with the result that variables that behave similarly appear closer to each other. Abbreviations: aggr, aggregation; ApoA-I, apolipoprotein A-I; BMI, body mass index; C, cholesterol; CETP, Cholesteryl ester transfer protein; CRP, C-reactive protein; HAE, HDL-apoA-I exchange; HDL, high-density lipoprotein; LCAT, Lecithin–cholesterol acyltransferase; LDL, low-density lipoprotein; PLTP, Phospholipid transfer protein; PON1, Paraoxonase and arylesterase 1; PP, per particle; TL, total load.

*Supplementary Figure 5*

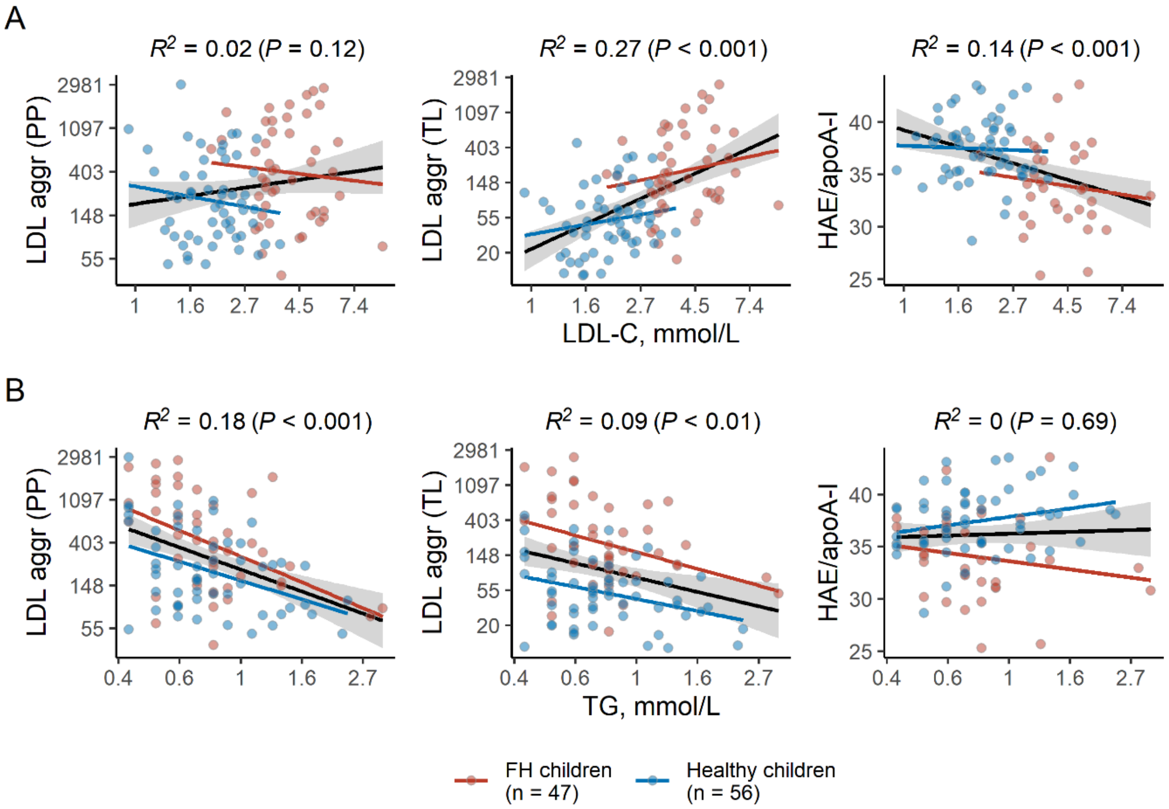


**Supplementary Figure 5**. *LDL-C and triglycerides associate with LDL aggregation and HAE/apoA-I ratio*. The figure shows the association between LDL-C (upper row: panel A) or triglycerides (TG, lower row: panel B) and measures of lipoprotein function (LDL aggregation (per particle or total load) or HAE/apoA-I ratio). Red and blue points represent FH children and healthy children, respectively, along with corresponding least-squares regression lines for those subgroups. The black regression lines and corresponding $R^2$ and P values are based on *all* children. Note that the axes are $\log_e$ transformed for LDL-C, TG and LDL aggregation (PP) and LDL aggregation (TL), but not for HAE/apoA-I ratio; this is in accordance with how these variables were treated in the analyses of the manuscript. For ease of interpretation, we labelled the axes with the familiar values (1-7.4 mmol/L for LDL-C and 0.4-2.7 mmol/L for TG). Abbreviations: aggr, aggregation; apoA-I, apolipoprotein A-I; C, cholesterol; HAE, HDL-apoA-I exchange; HDL, high-density lipoprotein; LDL, low-density lipoprotein; PP, per particle; $R^2$, r-squared (explained variance); TG, triglycerides; TL, total load.
